# Toward improving outcomes from early intervention for vulnerable children: pilot RCT of a new treatment for non-responders to parent training for conduct problems

**DOI:** 10.1101/2025.08.05.25332581

**Authors:** Helen Sharp, Hermione Roff, Nicky Wright, Andrew Pickles, Jonathan Hill

## Abstract

**Background:** Children with conduct problems are at high risk of a wide range of mental health problems in later life, making them a priority for early intervention. Group-based parent-training is known to be effective but with a substantial failure rate. Based on evidence on the value of involving children, we developed Reflective Interpersonal Therapy for Children and Parents, (RICAP). We report here, feasibility and outcomes from the first trial of an intervention for children with conduct problems persisting after parent training. In contrast to most other studies, we used both parent and teacher report.

**Methods:** The sample comprised 105 children and their parents aged 5-10 years referred to UK Child and Adolescent Mental Health Services (CAMHS) with conduct problems. All were offered the Incredible Years (IY) parent training intervention, and parents provided pre- and post-treatment measures (including CBCL, SDQ). Children still above clinical threshold after IY were randomized either to RICAP or to usual CAMHS treatment (CTAU) with follow up 8 months later. Trial Registration Number: ISRCTN25252940.

**Results:** Feasibility was supported by high retention through the initial IY (102/105) and subsequent RCT phases of the study (58/70 eligible for randomization). The majority of those randomized to RICAP attended for 11/14 or more sessions, reflecting its high acceptability to both children and parents. By parent report RICAP was superior to CTAU on CBCL externalising (d=0.32) and internalising (d=0.42) problems, while by teacher report CTAU was superior on SDQ total problems (d=0.32) and reactive aggression (d = 0.27).

**Conclusions:** We provide first evidence of the acceptability and effectiveness of a novel intervention for children with persisting conduct problem following parent training. We also find differences between parent and teacher reported outcomes, pointing either to reporter or social context effects, both of which need to be addressed in future research.

## Introduction

Disruptive behavioural disorders are among the most prevalent disorders in childhood, with 5% of British children meeting International Classification of Disease (ICD-10) criteria for conduct disorder and oppositional defiant disorder (ODD) [1] and global prevalence estimated to be 5.7% [2]. Multiple agencies are involved in responding to these difficulties, with a high proportion of children presenting to GP and emergency services regularly with conduct related issues [3]. Antisocial, disruptive or aggressive behaviour is a presenting problem in 30% of a typical GP’s child consultations and 45% of consultations in community child health [4]. In England, individuals with conduct disorder in childhood cost society ten times as much as control children by adulthood [3] and in the United States the lifetime cost to the public of conduct problems was estimated at $2.3 million [5].

Numerous longitudinal general population studies have confirmed that disruptive behavior problems seen in young children commonly persist, and that they are associated with increased risks not only for antisocial and poor interpersonal outcomes, but also for many psychiatric disorders, notably, depression, anxiety disorders, PTSD, and alcohol and drug misuse [6,7]. Equally, in spite of this markedly elevated risk, the majority of children with early onset disruptive behavior problems (estimated at between 50 – 70%) do not show persistence of those problems into adult life, although even those whose behaviours have improved have worse mental, physical, social and occupational outcomes compared to children with low lifetime problems [8,9].

For those with the ‘life-course persistent’ problems, outcomes are commonly very poor. In adolescence these children are at increased risk of poor peer relationships, exclusion from education, higher risk of unemployment, and involvement in criminal activity [10,11], depression [12,13] and suicidality [14]. In adulthood, poor outcomes include features of antisocial personality disorder such as criminality and poor interpersonal and occupational functioning, and the risks extend to a wide range of disorders, notably depression and drug and alcohol misuse, and to partner violence, negative parenting and physical abuse and hence risk to the next generation [7,8,15,16]. Children of antisocial adults are more often exposed to harsh and inconsistent parenting, low warmth, and reduced parental sensitivity, each of which is strongly associated with the onset and maintenance of child behavioural problems [17–19].

The UK National Institute for Clinical Excellence [4] recommend the use of manualised group-based parent training interventions such as the Incredible Years (IY) video-based parent training programme [20] as a first line treatment response for individuals referred with ODD or conduct problems. Informed by behavioural and social learning theories, such interventions aim to address maladaptive patterns of reinforcement of unwanted behaviours and are efficacious in specialist research settings [21,22], in ‘real world’ child and adolescent mental health services [23] and in Sure Start or voluntary sector settings to prevent conduct problems in preschool children [24,25]. However, a sizeable proportion of families drop-out prematurely or do not attend group interventions (30-50%) and a substantial number of children remain in the clinical range following treatment [3,26–28]. A major concern is that factors known to predict poorer outcome in parent training also predict persistence of antisocial behaviour in longitudinal studies; socio-economic deprivation, severity of child conduct problems, parental mental health problems, parental unresolved loss or trauma and partner violence in the home [1,29–32]. So, a serious challenge remains to develop interventions that address the needs of these families. Scott and Dadds [28] propose that a multi-model approach incorporating strategies based on alternative theoretical approaches such as attachment theory, structural family systems theory and cognitive-attribution theory may enhance the effectiveness of parent training.

Currently there are no evidence-based interventions for conduct problems which have been unresponsive to parent training. In the context of how to help where parent training has not led to substantial improvement, one of the recurring views expressed by parents is that the particular needs of their children have not been addressed [33]. Furthermore, approaches that do not include the child do not target drivers of disruptive behaviour problems which are internal to the child, nor do they take account of their individual differences. Addressing individual child vulnerabilities may therefore be key.

An extensive research literature shows that individual child vulnerabilities are strongly associated with persistent forms of antisocial behaviour and these characteristics may also limit the success of parent-focussed interventions. Such vulnerabilities include limited verbal skills, pragmatic language problems, deficits in executive function and information processing biases contributing to misperception of hostile cues from others and to inflated self-appraisal of competence [34–36]. Insecure parent-child attachment relationships are also significantly over-represented, and these are associated with child difficulties in interpreting and responding to other people’s emotions particularly when under conditions of emotional threat [10,37]. Another set of vulnerabilities characterise a distinct group of children with conduct problems who also show callous-unemotional (CU) traits (i.e a lack of guilt, empathy and concern for others, unconcern about their performance and shallow affect [38]). Children with CU traits experience greater conduct problem severity and higher absolute rates of symptoms after treatment compared to children without CU traits [39].

There are therefore good reasons to include children in interventions designed to improve outcomes for those not substantially helped by parent training. Several child focussed approaches have been developed and evaluated. Social skills training for children with disruptive behaviour problems has been shown to be effective, albeit with small effects [40]. The Coping Power Program (CPP) combines work with the child and parent training. The child approach uses a cognitive-behavioural framework to enhance the child’s ability to cope adaptively with difficult situations and feelings [41]. These interventions were originally designed to prevent the development of antisocial outcomes in children at risk, but more recently have been adapted to clinical samples in Sweden, Netherlands and Italy [42–44]. There is evidence that the effectiveness of CPP is sustained over up to six years [45]. However, to our knowledge, neither of these approaches has been evaluated as treatment for parent-training non-responders.

In designing the intervention for this study, Reflective Interpersonal Therapy for Children and Parents (RICAP), we took account of the considerable heterogeneity within the conduct disorders [10, 46–47]. This has been characterised in several ways, but a prevailing theme across several proposed typologies is the contrast between reactive anger prone opposition, and proactive more emotionally neutral or cold, low empathy, disruption or aggression [48–52]. We also took account of the possible role of anxiety in driving some oppositional or aggressive behaviours. While evidence for this is limited, a key hypothesis for the developmental consequences of disorganized attachment is that the child creates their own organisation through controlling behaviour, and importantly for RICAP ‘controlling-punitive’ behaviours [53–54]. The controlling behaviours are thus an attempt to deal with anxieties for which the child does not turn to the parent. Crucially, the insecurity and anxieties may not be apparent to the caregiver, because only the disruptive behaviours are apparent. RICAP therefore aims to create an arena in which the child’s anxieties can be identified and understood by the parent leading to changes in parental behaviour.

Enhancing a child’s capacity to understand and reflect on their own and other’s thoughts and feelings was in our view also a candidate for the focus of intervention in all of these cases. Increased reflectiveness would create opportunities for children to modify social cognitions associated with anger proneness [34] to respond empathetically to others, and also to recognise their own underlying feelings, learn new ways of expressing them and to make new choices for interpersonal behaviour as a result. Finally, the development of RICAP was also informed by findings from our research group, using story stems in the context of doll play. In response to emotionally challenging doll-play scenarios, children who showed low ‘intentionality’ on a scale assessing the extent to which the motives of the doll participants were described or demonstrated in their play had higher levels of disruptive behaviour problems [37,55] further suggesting that an intervention designed to increase child reflective functioning within interpersonal exchanges may be helpful.

The RICAP method contains some structure (e.g., certain topics for drawing are introduced) but it is flexible enough to ensure a personalised approach. Table 1 gives a summary of the aims and components within the intervention. The approach with the child makes use of both the representational content and process of child drawing to help identify attachment related concerns and social-cognitive or attributional biases in the child’s view of the world. The therapist-child conversation in session is designed to promote an autonomous capacity to reflect and self-reflect, to broaden the child’s understanding of self and others in interpersonal exchanges. This in turn opens up new possibilities for adaptive emotional expression and alternative child behavioural choices. A book is compiled by the child therapist to record the drawings and conversations from each session. Since many of these difficulties are manifested within the parent-child relationship, we developed a parallel approach with parents designed to increase their own reflective capacity about self and other, including understanding the range of underlying feelings and motivations that might underpin their child’s opposition, defiance or aggression in specific circumstances that occur day to day. The method uses curious questioning to tease apart in detail the sequence or chain of events that occurred in the parent-child interactions during a good time and difficult time which the parent chooses from the past week. The aim was to help parents build new understandings of their child and generate a broader repertoire of possible responses in the light of those understandings, so they might feel able to make more informed choices about how to respond in a given situation. These new understandings then have the potential to increase parents’ ability listen to their child, to respond empathically and consistently [56,57] whilst also setting appropriate behavioural limits when required.

**Table 1.**
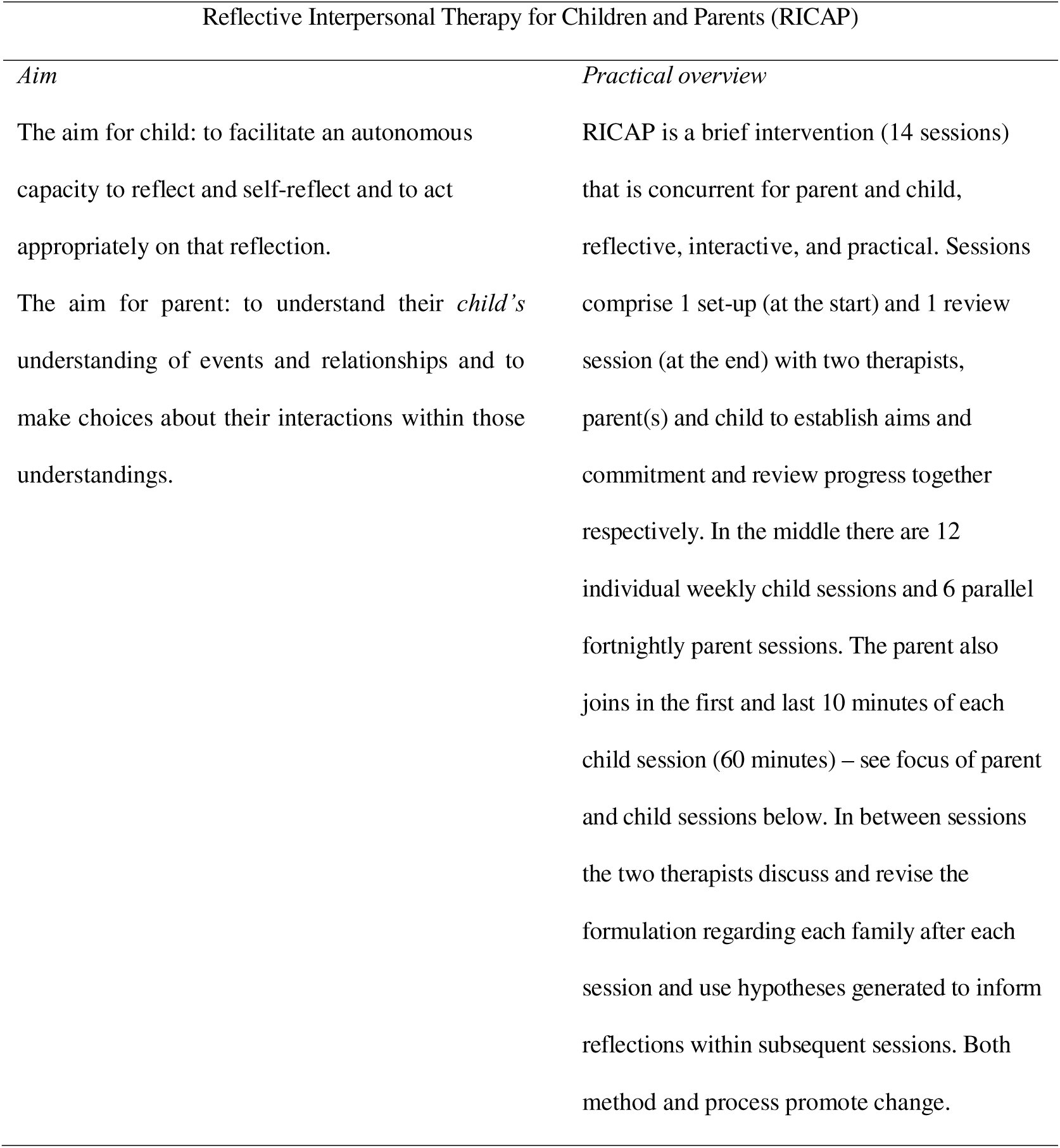
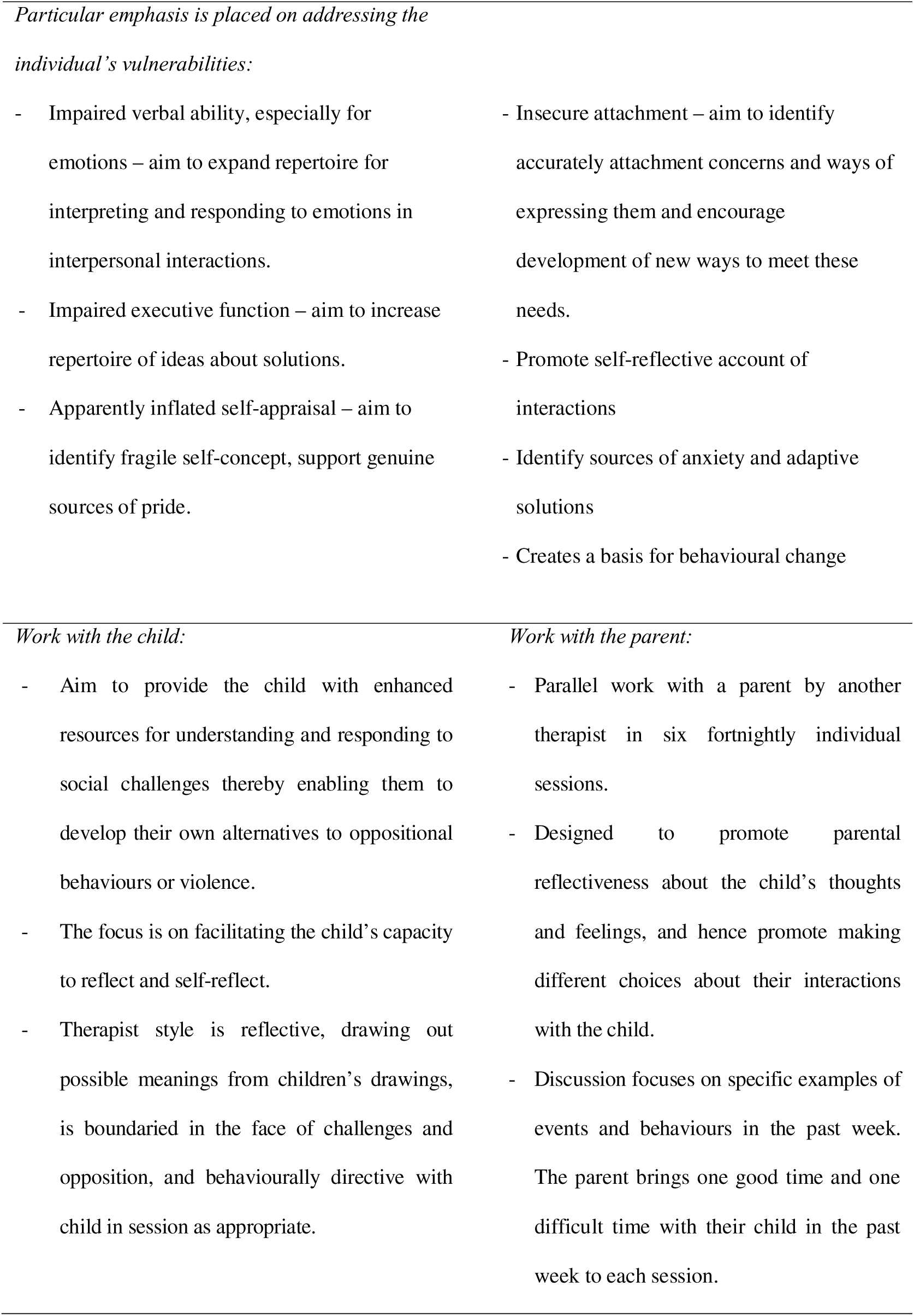
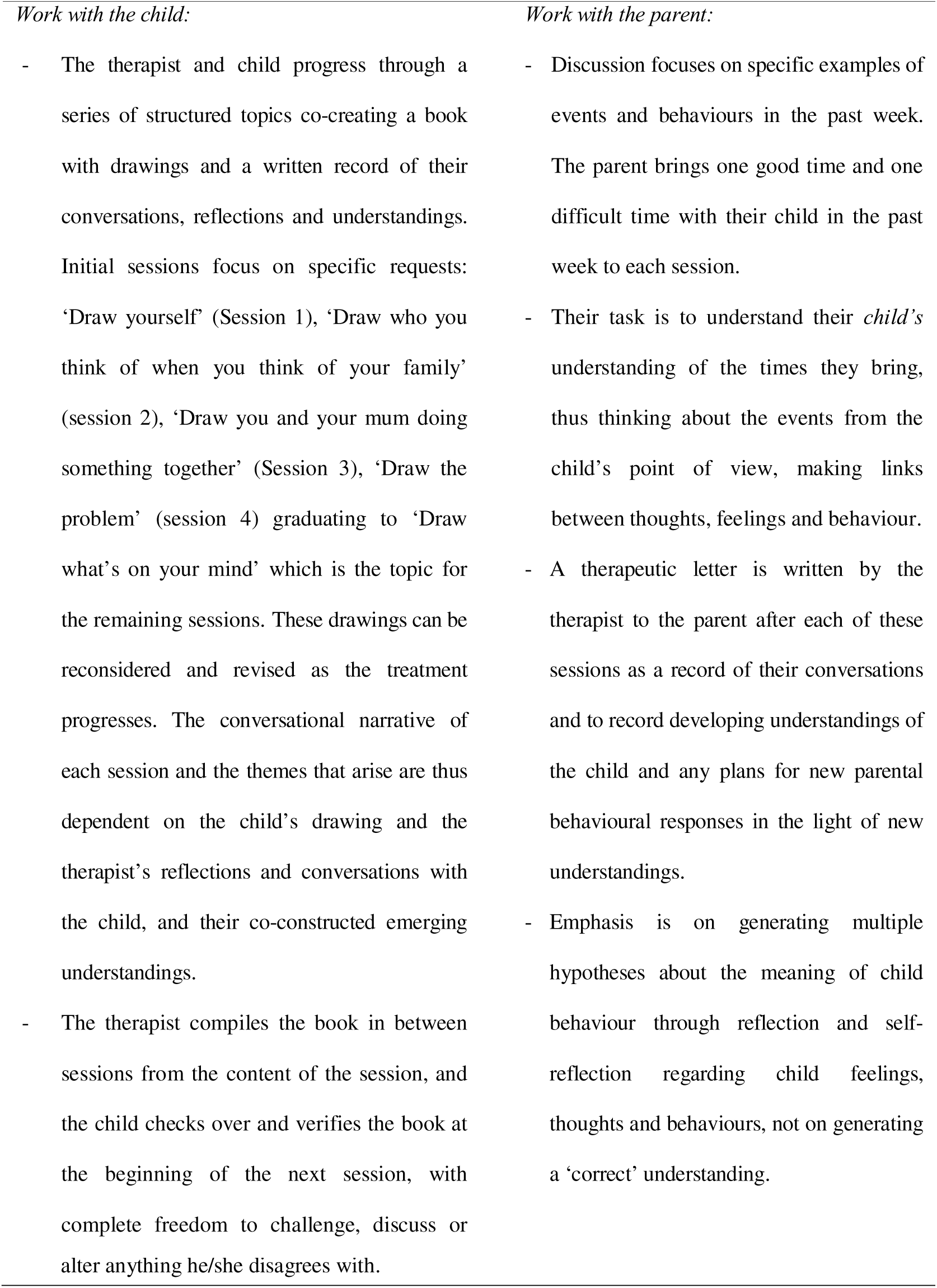
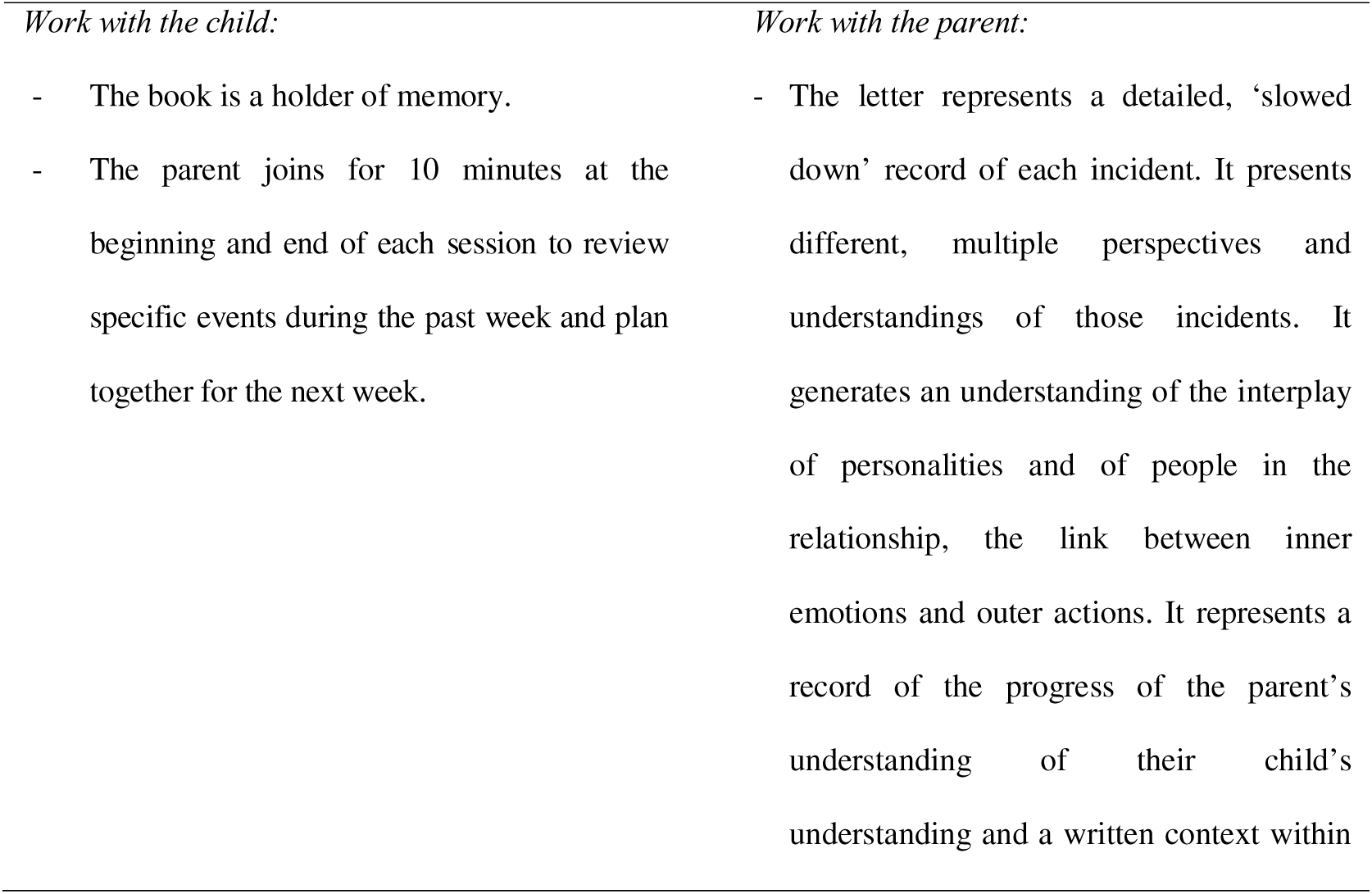
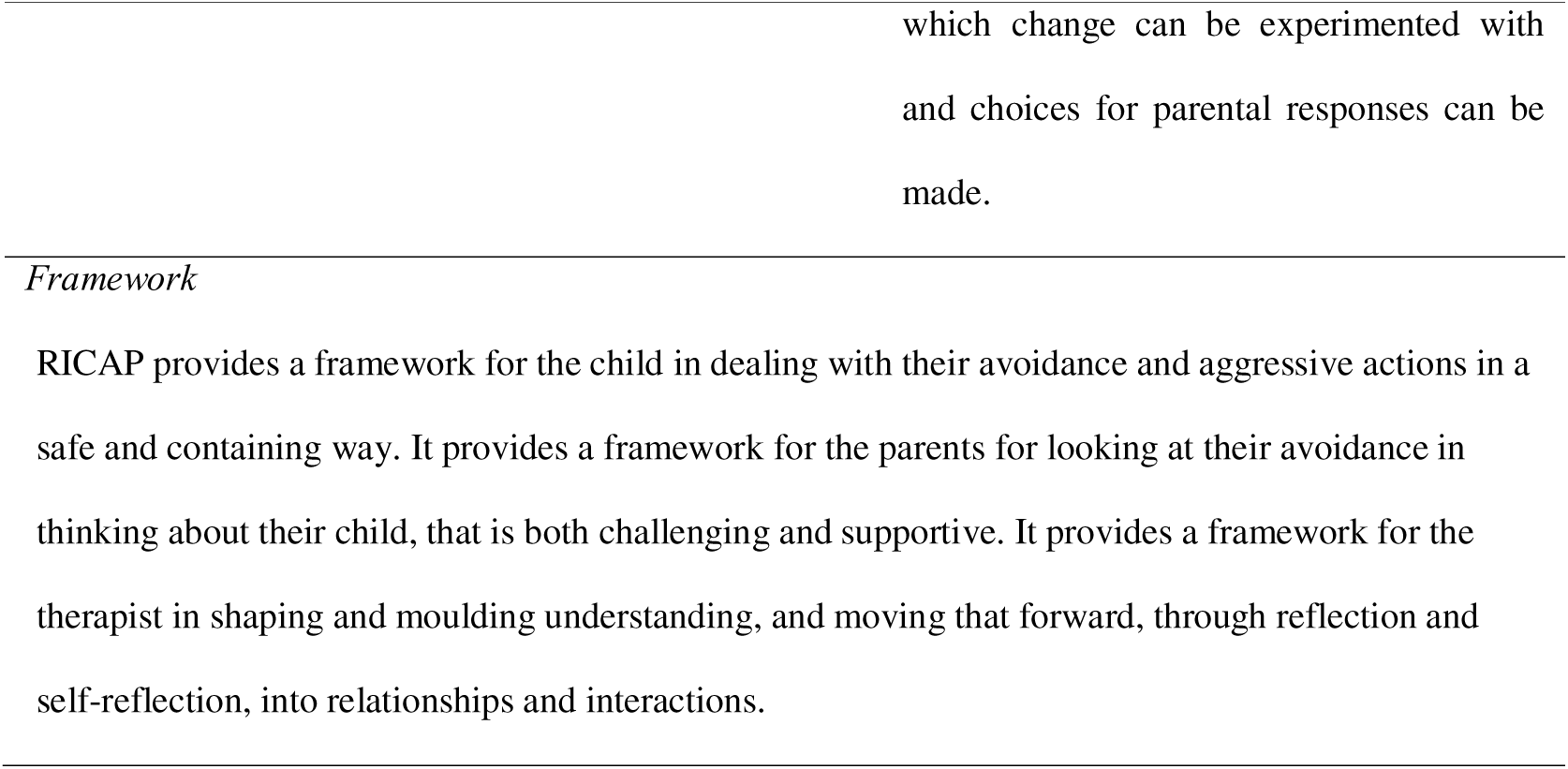
Reflective Interpersonal Therapy for Children and Parents (RICAP) description.

The overarching logic model for change in RICAP includes both child and parent processes. The child’s increased awareness of their emotional states and thoughts provide them with alternative coping possibilities and facilitates more accurate communication of their needs to parents. The child can also make new behavioural choices in the light of their own new understandings. More accurate communication to parents in turn makes it more possible for a parent to match their responses to the child’s needs. The parent’s increased reflectiveness enables them to go beyond the immediate behaviours of opposition and aggression, to consider a range of alternative drivers and hence broaden their repertoire of responses. Thus the parent is able to refine their responses, for example responding firmly when guidance is required, and tenderly where comfort and understanding are needed. Change also follows from the parent’s effective and accurate responding in these different domains of parent-child interaction.

In summary, ‘Reflective Interpersonal Therapy for Children and Parents’ (RICAP [56]) is a standardised and at the same time flexible intervention for conduct problems designed to address individual child and parent vulnerabilities. RICAP aims to enhance children and parents’ awareness of thoughts and feelings predominantly in the parent-child relationship, in order to provide a platform for alternative less conflictual or hostile interactions.

We report results from a pragmatic pilot randomised controlled trial (RCT) in which children rated in the clinical range for conduct problems either by parents or by teacher at the end of a ‘gold standard’ parent training intervention (The IY programme [20]) were randomised to receive RICAP or clinical treatment-as-usual (CTAU). There were three main aims of the study. First to assess feasibility of the design within UK NHS settings, and in particular to determine retention across a study with two phases, first of IY intervention, and second randomization of children with persisting behaviour problems. Second, to find out whether families not helped by previous intervention took up, and stayed in, the new treatment, RICAP. Finally our aim was to generate effect sizes from a range of outcome measures in order to inform future full trial design.

## Methods

### Study population

Eligible children were aged 4.0-11.0 years referred for child conduct problems to Child and Adolescent Mental Health services (CAMHS) or multiagency community behavioural services (Behaviour Education Support Teams) on Merseyside, UK. These services were identified because they had experienced therapists routinely offering the IY Programme to groups of parents as a first line intervention. Participants were required to be English speaking.

The criterion for inclusion in the study was a score in the clinical range on either the CBCL/TRF externalising subscale or SDQ conduct problems subscale from parent and/or teacher informant (see Measures). Children with a prior diagnosis of ADHD were eligible if not receiving medication or if on an established dose of stimulant. Ethical approvals were gained from the Liverpool Children’s Research Ethics Committee and Wirral Health Authority Research Ethics Committee (REF 03/05/051/C) and written informed consent was obtained from all parents. The sample size calculation is shown in Supplementary Materials 1.

### Design

A two-phase pragmatic pilot RCT (Trial Registration Number ISRCTN25252940 https://www.isrctn.com/ISRCTN25252940). The trial was not registered prior to the start of recruitment because at that time, September 2003, the International Committee of Medical Journal Editors (ICJME) requirement for trial registration had not been introduced.

In Phase 1 all parents who consented to take part in the study, and met the inclusion criterion were offered group-based IY parent-training. Following the Phase 1 parenting-training, at 4 months, families with children rated in the clinical range either on parent or teacher reported CBCL, TRF or SDQ were eligible for randomization either to RICAP or CTAU in Phase 2.

Randomization was by made concealed allocation. A block randomisation design was used, using blocks of six to ensure similar sample sizes in each arm. The researcher made a telephone call to an office where the randomisation schedule, devised by an independent statistician, was available to a secretary with no knowledge of the trial participants who provided the allocation. Measures were administered prior to Phase 1 (Baseline, Time 1), 4 months later (time 2 – post phase 1 treatment and pre phase 2 treatment) and 12-14 months after baseline (time 3-post phase 2 treatment). Time 3 measures were gathered by research assistants who had not carried out previous assessments with the family, blind to treatment allocation. Teachers were not informed of the children’s treatment allocation. All families were followed up at one year after initial recruitment regardless of whether they had received a phase 2 intervention or not. At the point of consent parents were informed that the study was designed to compare ‘two different ways of working with parents and children’ after parent training. They were not informed that one was a novel therapeutic approach so as to minimise expectancy effects. The instructions given to participants are shown in Supplementary Materials 2. Recruitment began on 04/09/2003 and the trial ended on 21/09/2006.

The study was designed according to the MRC guidelines on Phase Two Exploratory Studies of complex interventions (MRC, 2000) and the CONSORT statement [58,59]. The trial details are reported in line with the CONSORT guidelines for reporting of parallel, pragmatic and non-pharmacological interventions [60].

### Measures

#### Child Behaviours

Child Behaviour Checklist (CBCL [61]) and teacher report Teacher Report Form (TRF [62]) to assess externalising, internalising and total problems at Time 1, Time 2, and Time 3 (parent report α=.81; teacher report α=.85), Time 2 (parent report α=.82; teacher report α=.84) and Time 3 (parent report α=87.; teacher report α=.85).

Strengths and Difficulties Questionnaire (SDQ [63]) parent and teacher report to assess conduct problems and total problems, at Time 1 (parent report α=.69; teacher report α=.75), Time 2 (parent report α=.74; teacher report α=.77) and Time 3 (parent report α=72.; teacher report α=.78).

Parent Daily Report (PDR [64]) records 31 problem behaviours as present or absent each day for up to 7 days and was administered over the telephone over a 1-2 week period, at Time 2, and Time 3.

Teacher report of Reactive and Proactive Aggression [50] at Time 2 and Time 3.

### Parenting practices

Frequency of smacking was recorded using the Parent Daily Report at Times 2 and 3.

### Main carer mental health

Twenty-eight item General Health Questionnaire (GHQ-28) [65] at Times 1, 2 and 3.

### Demographic information

Main carer and child ages, marital/partner status, age carer left full time education, eligibility for free school meals, and ethnicity were all recorded. The index of multiple deprivation, based on postcode was calculated for each household [66]. A binary variable reflecting presence or absence of the most deprived quintile was used.

### Interventions

Phase 1 intervention was the IY videotape parent management training (PMT) programme [20] delivered over 12-14 weeks according to the manual and in accordance with NICE (2006) guidelines [67]. All PMT therapists had completed an accredited IY training course.

Phase 2 interventions comprised RICAP and CTAU. RICAP was developed to further meet the individual treatment needs of these families by Hermione Roff working within the academic group at the University of Liverpool. As outlined earlier, RICAP is a brief 14 session manualised therapy which involves two therapists working in parallel with parent(s)/carer(s) and child. It is aimed at children aged 4-10 years of age. Table 1 gives a summary of the aims and components within the intervention. See Roff [56] for a full description of the approach.

RICAP was delivered by experienced CAMHS therapists across eight patch teams who were newly trained in the approach and who had completed two supervised cases prior to working with families on the trial. Treatment integrity was ensured and monitored in a number of ways; (a) three days of training using a manualised description of the key components of the therapy and illustration with case examples (b) fortnightly supervision during completion of practice cases and study cases by the originator of the therapy (c) examination of adherence to the therapeutic model through the content within the child’s book of in-session drawings and conversation, and the therapeutic letters sent to the parent. The intervention was typically delivered over a 16 week time period.

CTAU comprised treatments offered routinely in the clinical settings, including individual parent behaviour management interventions, school based intervention, family therapy, child cognitive behavioural therapy, anger management training, child psychotherapy, problem-solving skills training, supportive advice and listening, psychopharmacological intervention for ADHD, or a combination of these elements. Of those randomized to CTAU, 42% of the children received a school-based intervention component through Behavioural Education Support Teams (BEST) as an additional component of their CTAU. Examples of school-based intervention included restorative justice, emotional literacy, educational psychology involvement and broadening supports with school-based activities. The majority of families received two or three co-occurring clinical interventions typically parent and child and/or school focussed (63%). The most common clinical components of CTAU were individualised parent behaviour management training, ADHD diagnosis and medication, and anger management training.

### Analysis plan

Feasibility was established by observing rates of parental attendance. Skewed variables were transformed used square root transformation. Paired t-tests were used to determine the effect of phase 1 treatment. Repeated-measures Analysis of Covariance (ANCOVA) was used to test for effect of treatment allocation during phase 2 on post-intervention scores, after covarying for the possible effects of baseline child behaviour, child age, sex and deprivation [68]. Intention to treat analyses is also reported, assuming no change in scores for those cases lost to time 3 follow-up but who gave valid time 2 data. We report analyses with all allocated cases with complete follow up data, regardless of how many intervention sessions they received. Clinically reliable change using the borderline and clinical cut offs on the CBCL and SDQ is reported.

## Results

### Recruitment and retention, feasibility

Recruitment and retention rates were used to assess feasibility. Recruitment to Phase 1 treatment is shown in Figure 1.

**Figure 1.**
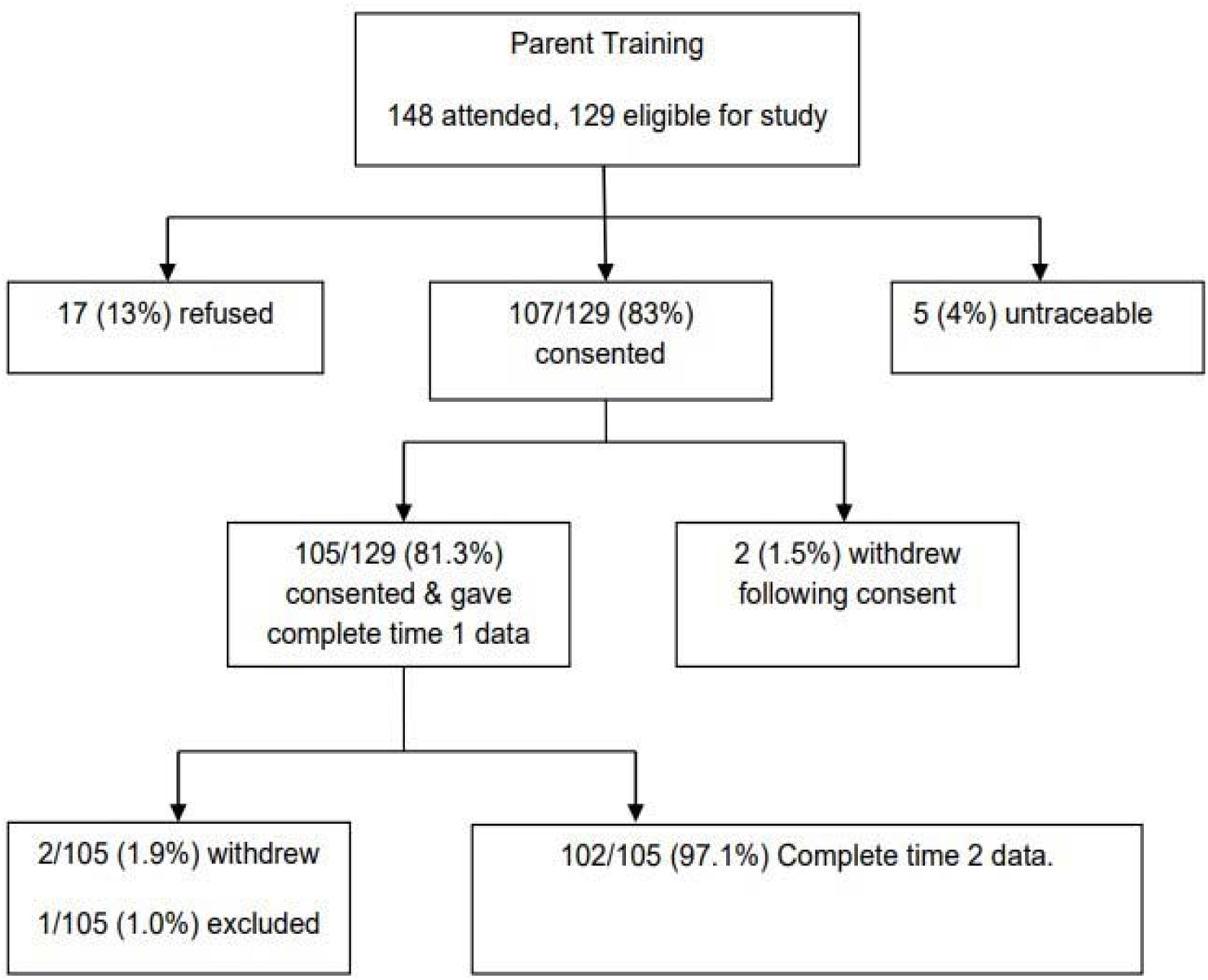
Recruitment to Phase 1 parent training (IY) for all families

Numbers eligible for Phase 2 because above clinical threshold on either parent or teacher report are shown in Figure 2.

**Figure 2.**
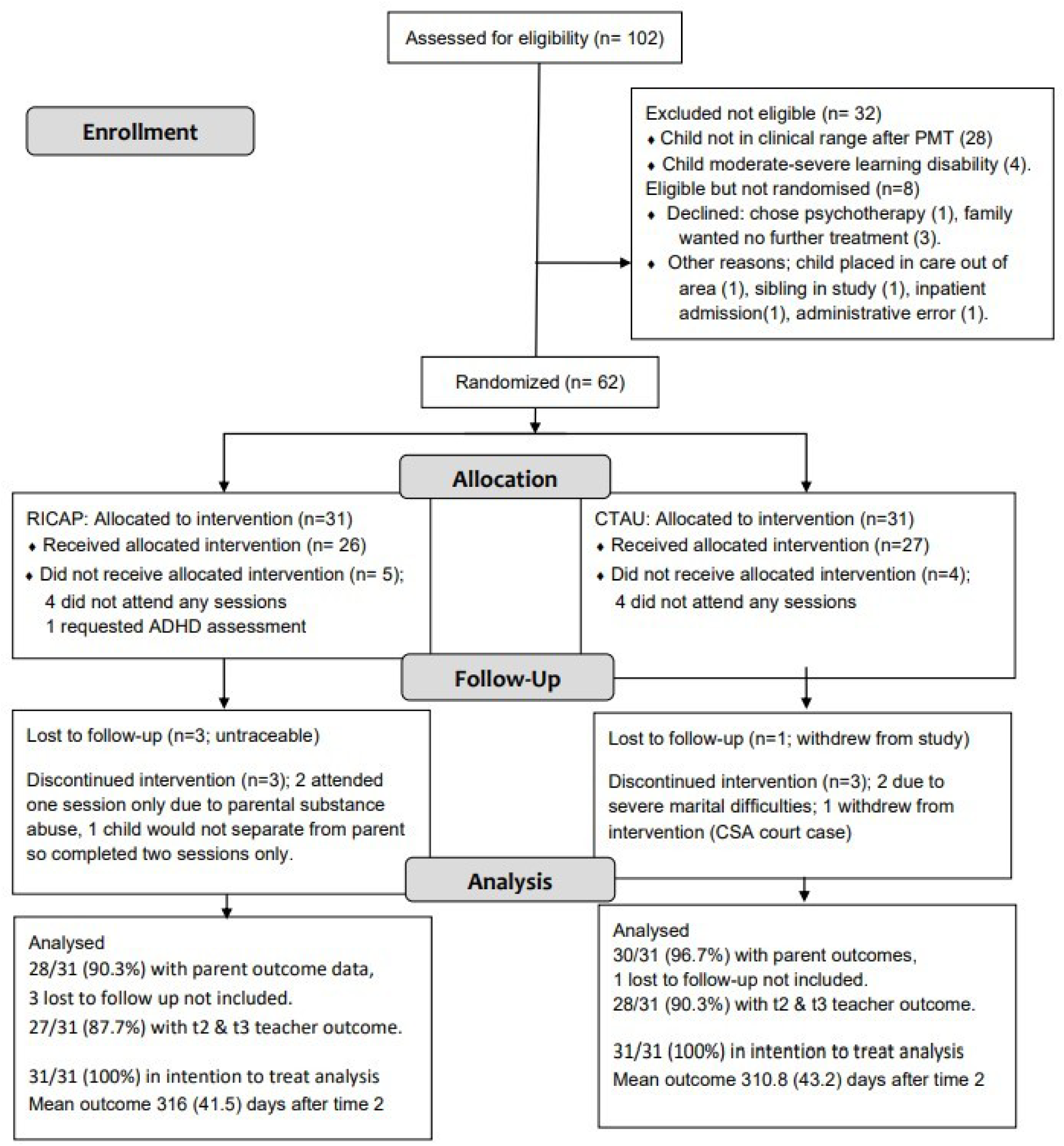
Recruitment to Phase 2 randomization to either RICAP or CTAU.

A high proportion of parents (102/105; 97.1%) and teachers (94/102; 92.2%) completed measures before and after IY treatment. There was a total of 72/102 (72.5%) children above the clinical threshold after parent training, two of whom had moderate to severe learning disability leaving 70 eligible for phase 2. One family allocated to RICAP received CTAU after requesting an ADHD assessment, and no family allocated CTAU received RICAP. Completion rates in each arm of the trial were high and similar (28/31, 90.3%, for RICAP and 30/31, 96.8%, for CTAU).

The demographic characteristics of the sample recruited to Phase 1 are shown in Table 2. The children came from families with high levels of deprivation, single parenthood, and low levels of parental education. Just under two thirds of the sample were eligible for free school meals. Only one family was not white British, reflecting the low level of ethnic minorities in the areas of Liverpool and the Wirral. Randomisation yielded two groups of families for Phase 2 intervention with similar profiles. The two groups did not differ in child age (*p* >0.05) or the percentage of PMT sessions attended in Phase 1 (*p*>0.05). They were also comparable in demographic composition and level of child antisocial behaviour symptoms on which randomisation was based (all p>0.10).

**Table 2.** Characteristics of children and their families referred for parent management training for antisocial behaviour (Phase 1 treatment); whole sample and subgroups defined by status at start of phase 2 treatment.

|  | Phase 1 | Baseline characteristics for Phase 2 subgroups |  |  |  | UK<br>mean □ |
| --- | --- | --- | --- | --- | --- | --- |
|  | Whole sample<br>N=102 | Non-clinical after<br>Phase 1 treatment<br>n=28 | Not randomised<br>n=12 | Randomised to<br>RICAP<br>n=31 | Randomised<br>to TAU<br>n=31 |  |
| Child male, n (%) | 67 (65.7) | 14 (50.0) | 8 (66.7) | 24 (77.4) | 21 (67.7) | - |
| Child mean age in months<br>(SD) | 90.16 (22.0) | 86.00 (21.4) | 90.58 (22.9) | 90.19 (22.0) | 93.71 (22.5) | - |
| Main carer age in years (SD) | 35.4 (7.35) | 33.2 (5.78) | 44.2 (9.37) | 33.4 (6.89) | 36.0 (5.87) | - |
| Mother left education by 16<br>years | 70 (68.5%) | 19 (67.8%) | 9 (75.0%) | 24 (77.4%) | 18 (58.1%) | 13% |
| Lone parent family | 44 (43.1%) | 11 (39.3%) | 6 (50.0%) | 13 (41.9%) | 14 (45.2%) | 7% |
| Most deprived UK IMD | 71 (69.6%) | 20 (71.4%) | 9 (75.0%) | 22 (71.0%) | 20 (64.5%) | 20% |

**Table 2** (continued).
|  | Phase 1 | Baseline characteristics for Phase 2 subgroups |  |  |  | UK<br>mean<sup>□</sup> |
| --- | --- | --- | --- | --- | --- | --- |
|  | Whole sample<br>N=102 | Non-clinical after<br>Phase 1 treatment<br>n=28 | Not randomised<br>n=12 | Randomised to<br>RICAP<br>n=31 | Randomised<br>to TAU<br>n=31 |  |
| No car | 46 (45.1%) | 12 (38.7%) | 7 (58.3%) | 14 (45.2%) | 12 (38.7%) | 28% |
| Ethnicity white British | 101 (99.0%) | 30 (96.8%) | 12 (100.0%) | 31 (100.0%) | 30 (96.8%) | 9% |
| Council / housing association<br>home | 49 (48.1%) | 9 (32.2%) | 8 (66.7%) | 16 (51.6%) | 16 (51.6%) | 17% |

<sup>□</sup> Data from *Social Trends* London: Office of National Statistics, 2000.

### Phase 1, Parent Management Training Outcomes

Mean attendance for the PMT groups in Phase 1 was 7.41 (SD 4.6) sessions. There was a substantial decrease in parent-reported CBCL externalising (*d* = 0.73) and SDQ conduct problems (*d*=0.68) and a small decrease on teacher-report TRF externalising (*d*=0.27) and SDQ conduct problems (*d*=0.22) from Time 1 to Time 2. The size of the treatment effects derived from the two parent outcome measures were similar in magnitude to those reported by Scott et al. (2001) in a UK community RCT comparing the effectiveness of PMT with wait-list controls in clinical settings. Table S1 gives the means and standard deviations pre-and post-PMT intervention for the whole sample, and also subgroups defined by their status at the point of random allocation to phase 2 intervention.

### Phase 2, Participation in RICAP

Figure 3 shows the number of sessions of RICAP attended by families expressed as a function of the level of parent’s attendance during phase 1 PMT intervention. Since parent and child sessions occur on the same occasion, the maximum is 14 sessions together. Of the 31 who were randomised to receive RICAP, 23 (74%) attended for six sessions or more, and 5 (16.1%) did not attend any sessions of RICAP. Importantly there were 13 families in the RICAP group where parents had attended four or fewer IY PMT groups (defined in Scott et al [3]) as drop-out), and of these seven attended eleven or more RICAP sessions. Within the CTAU arm 4/31 (12.9%) families did not attend any CTAU sessions. Adherence to CTAU could not be further determined as interventions varied and clinicians did not all define a set length of treatment at the start. Of note is the fact that 13/31 (42%) of CTAU recipients had an intervention that included a focus in both home and school settings, whereas RICAP recipients did not receive any school-based intervention.

**Figure 3.**
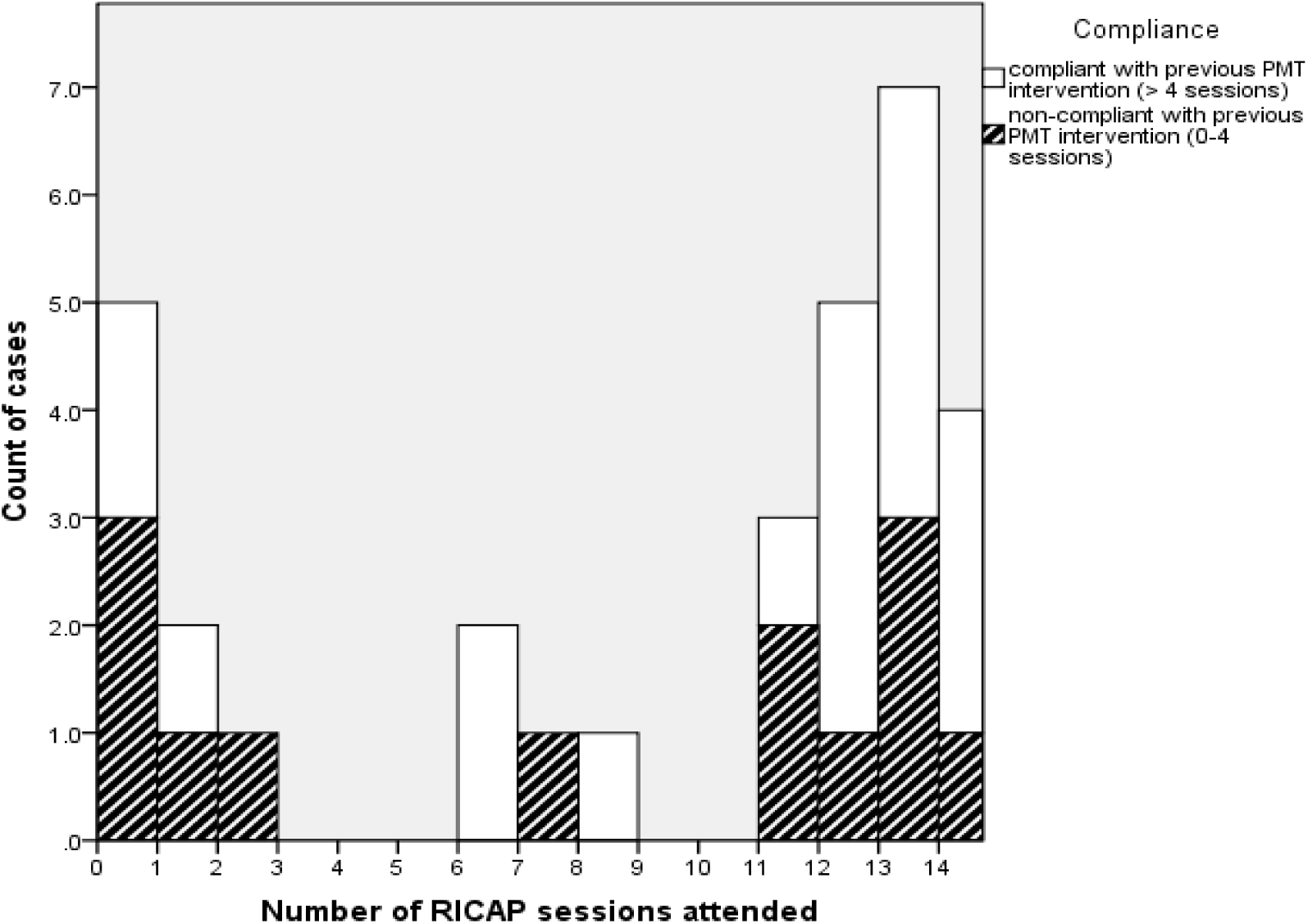
Participation in RICAP comparing families who attended five or more IY groups and those who attended no more than four.

### Phase 2 pilot RCT, child behaviour outcomes by parent report

The mean parent report scores at time 2 and time 3 with associated effect sizes are shown in Table 3. Analyses revealed significant small to medium treatment benefits associated with allocation to RICAP intervention on externalising problems (*d*=.37), internalising problems (*d*=.46) and on total child problems reported on the CBCL (*d*=.43). These benefits were reduced by 9 - 16 % in intention to treat analyses but remained small to moderate and significant. These effects were not seen with parent-report SDQ nor Parent Daily Report measures.

**Table 3.**
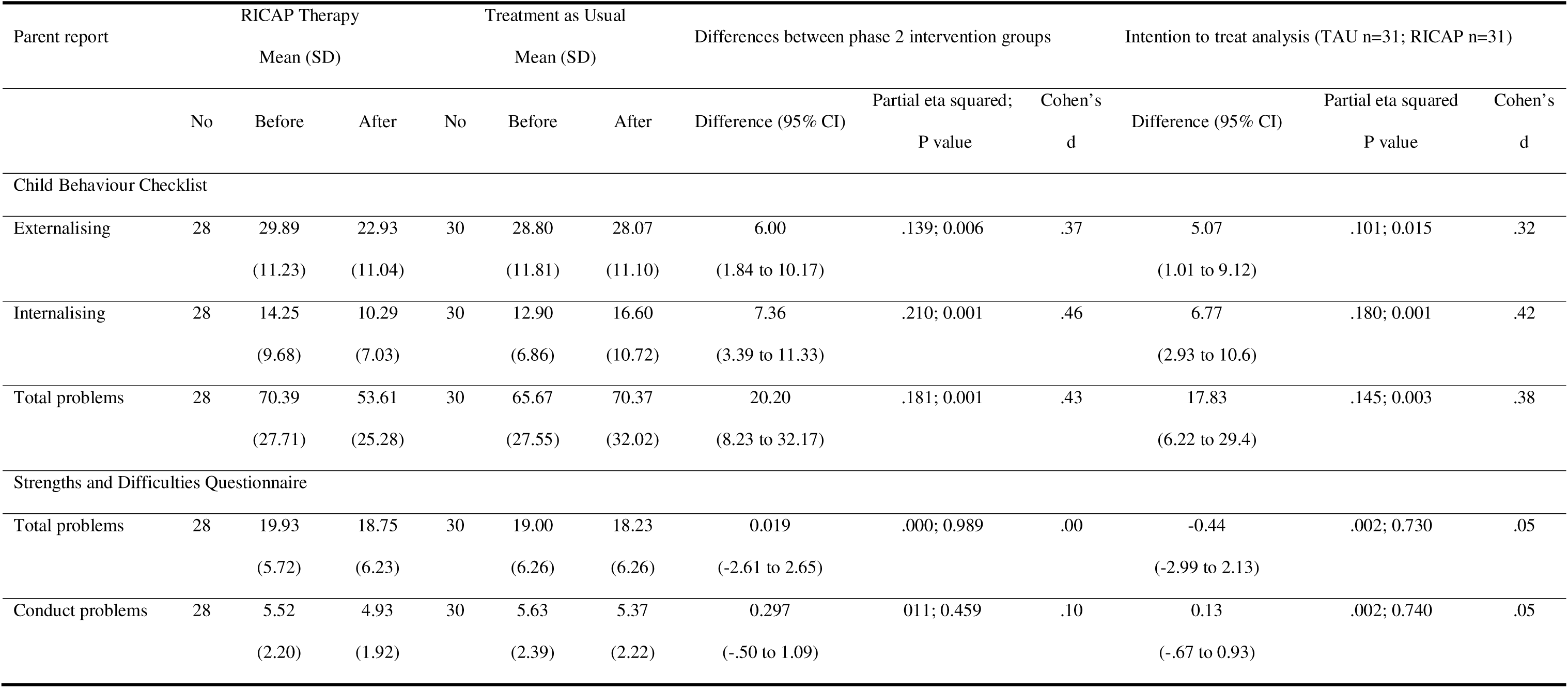
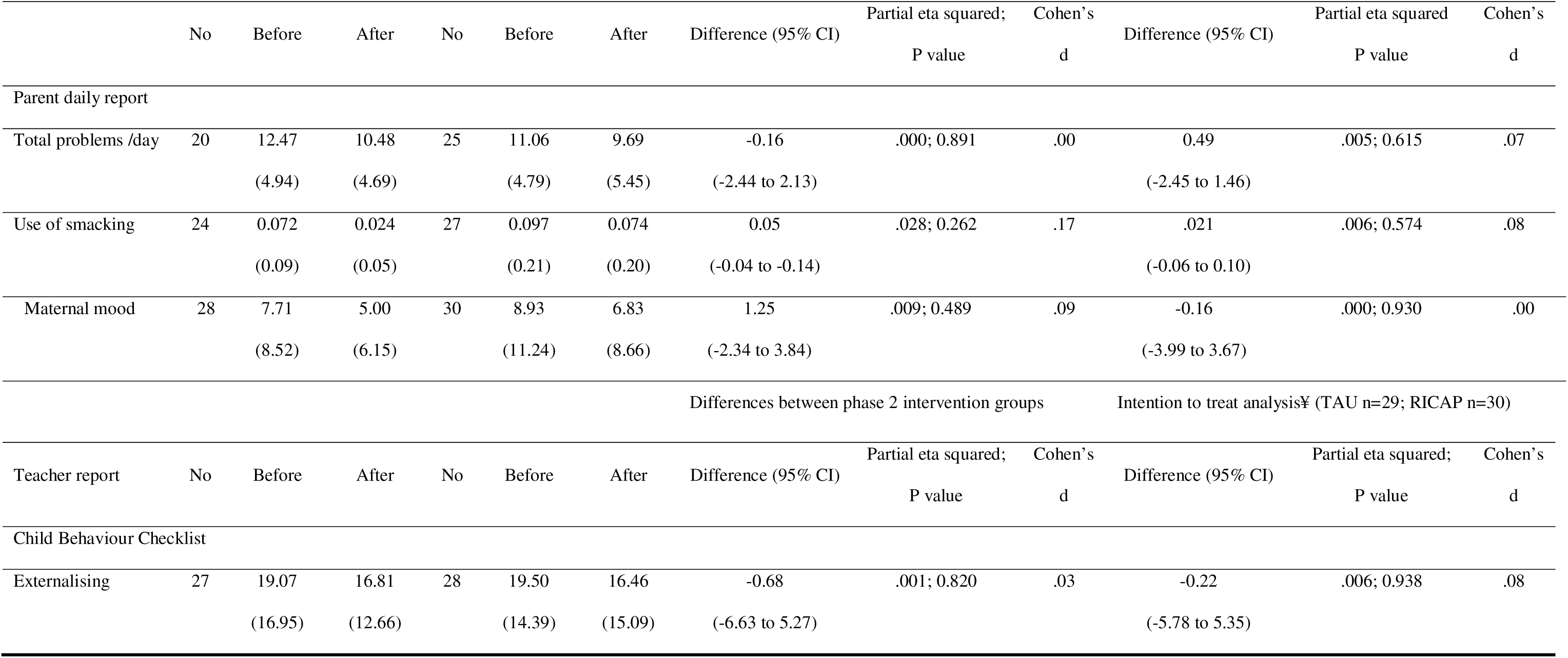
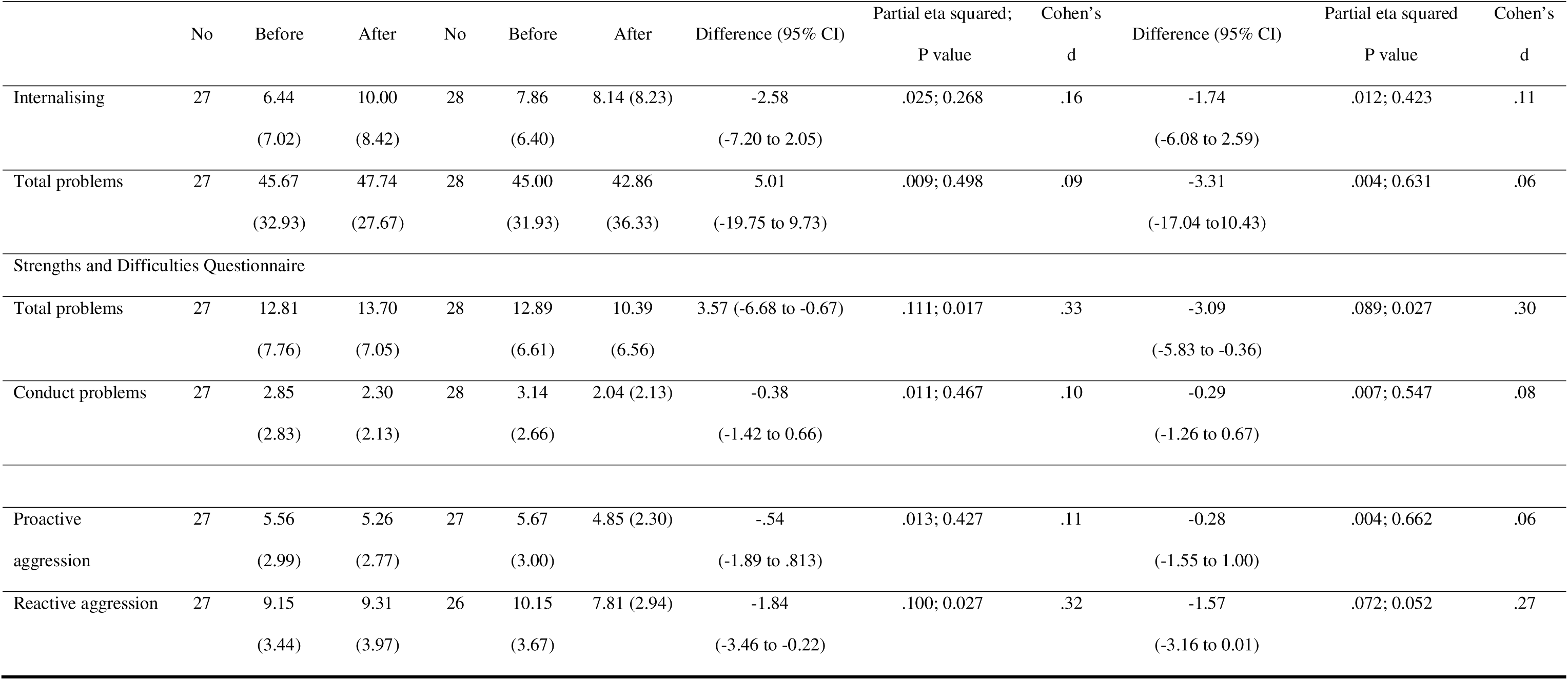
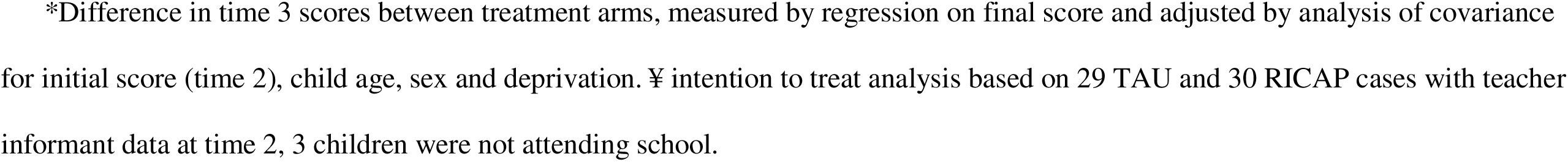
Summary of parent and teacher raw report scores before and after phase 2 intervention with per protocol and intention to treat effect sizes based on adjusted scores.

#### Clinically reliable change

Table S2 shows the percentage of participants rated within the normal, borderline or clinical ranges on the parent-report CBCL and SDQ. The proportion of children rated in the clinical range for CBCL externalising fell by 32% in the RICAP arm, and by only 12% in the CTAU arm, leaving 57% and 75% still in the clinical range at the end of intervention respectively. The proportion rated in the clinical range for internalising problems reduced by 27% in the RICAP arm, and increased by 15% in the CTAU arm, leaving 21.4% and 51.7% still in the clinical range at the end of intervention respectively. Clinically reliable change according to the SDQ measure did not differ between treatment arms.

### Phase 2, Outcomes from Teacher Reports

In contrast to parent reports, no group differences were observed in teacher rated reports in favour of RICAP. No group differences were observed on either the CBCL scales or the SDQ conduct problems subscale. Teachers reported lower levels of reactive aggression at time 3 in the CTAU group compared to the RICAP group. No differences were reported in proactive aggression. Teachers reported significantly lower levels of SDQ total problems at time 3 in the CTAU group. Intention to treat analysis revealed a similar pattern of results.

#### Clinically reliable change

The proportion of children rated in the clinical range within the school setting was observed to be lower than that reported in the home setting as a whole, but there were no differences between the RICAP and CTAU groups.

## Discussion

This pragmatic RCT confirmed previous evidence that many children do not improve with parent training. We demonstrated that we could retain these families in a second intervention, including many who dropped out of parent-training, and follow them up over a substantial period of time, with equal numbers retained in both arms of the intervention. We showed that UK NHS practitioners can be trained and supervised in this novel therapeutic approach. RICAP was found to have a clear advantage over CTAU for parent-reported problems on the CBCL in children who had not responded to parent-training. However, this was not the case for some teacher reported problems where there was evidence for superiority of the CTAU intervention.

Feasibility might be a concern for studies of conduct problems in children, because these are associated with socioeconomic deprivation [10], which in turn is associated with lower participation in medical research [69]. However the findings provide strong support the feasibility of conducting a two-phase trial of the treatment of conduct problems within the UK NHS. Notably there was a high level of retention of families over a period of at least a year, and a high completion rate of measures from parents and teachers. Of those recruited to the first, parent training, phase of the study, 97% provided follow up data, and of these, 89% consented to be randomized. 94% of those randomised provided follow up data. There was no evidence of differential attrition between the RICAP and CTAU groups.

As far as we are aware this is the first evaluation of an intervention designed to treat conduct problems which have not responded to ‘gold-standard’ parent-training. In the first phase of the study, parent-training was associated with moderate to large reductions in parent-reported conduct problems. Despite this reduction, 72.5% of children (72/102) remained in the clinical range according to parent or teacher report after parent-training and 70/72 were eligible for phase 2. Allocation to RICAP was associated with significant small-to-moderate reductions from post-phase 1 to post-phase 2 in parent-reported CBCL externalising, internalising and total problems. Allocation to RICAP was not associated with a significant reduction for parent-reported SDQ or daily report of problems compared to CTAU, and no significant treatment effects were observed for teacher ratings. The results provide initial evidence that RICAP delivered in regular clinical practice may be an effective and acceptable intervention for treatment-resistant conduct problems. This is encouraging when viewed in the context of the majority of studies of treatment for child conduct problems which only include parent reported outcomes. Teacher reported outcomes in this study provided a different picture, albeit on different measures. These indicated superiority of CAMHS based CTAU interventions,

Interpretation of the effect sizes from this trial should be considered in the context of several contextual factors. First, the sample represented complex cases who have not responded to parent-training and were characterised by high rates of risks for conduct problems including low parental education and deprivation. A greater proportion of the sample remained in the clinical range after parent-training compared to prior UK RCTs [3]. Second, this study was conducted with an unselected sample of referred children in usual clinical services and so is likely to be representative of children attending clinical services. Third, the trial was conducted by CAMHS practitioners after brief training in RICAP. They had typically only experienced being supervised with two practice cases prior to the trial proper. Therefore, the effectiveness estimates reported in this pragmatic trial may underestimate what might be achieved with more experienced RICAP therapists within clinical practice over time. In this context the small to moderate treatment gains in children allocated to RICAP provide promising initial evidence for effectiveness for treatment-resistant conduct problems. They were comparable to those reported in a meta-analysis of 52 evidence-based psychosocial interventions which were tested against usual clinical care [70].

UK NICE (2013) [4] guidelines acknowledge that complex families may need alternative approaches to first-line group-based parenting interventions devised to better meet their needs. However, rather than offer alternative approaches as a first line of approach, offering them once the outcomes from parent training is known, ensures both that everyone has had an evidence based intervention, and that they are being targeted to those most in need. The method and process of RICAP was devised specifically with the complexities of these families in mind. Whilst providing a structure for the therapists to work within, the considerable flexibility of formulation within the RICAP method allows different aspects of child and/or parent vulnerability to be addressed, arguably making it more sensitive to the heterogeneity evident in the most complex families. A substantial proportion of those families who did not adhere to the PMT intervention subsequently successfully adhered to RICAP (62%), indicating that the intervention was acceptable to these families. Unfortunately, it was not possible to examine comparable adherence to CTAU.

It is noteworthy that RICAP was equally as effective at reducing internalising problems as it was externalising problems. Recent meta-analyses of the effects of parent-training on internalising outcomes have concluded that there is evidence for a small effect but with significant heterogeneity across studies [71,72]. The focus in RICAP on children and parents’ awareness of thoughts and feelings and the targeting of anxieties which may underlie oppositional behaviour may mean that it is particularly suited to reducing co-occurring internalising problems. While RICAP treatment effects were observed for parent-reported CBCL externalising and internalising outcomes, there were no differences on parent-reported SDQ scores, on behaviours reported by parents by phone in the PDR, or teacher reported CBCL scores. Furthermore the CTAU group showed greater improvement than the RICAP group on teacher rated total (but not conduct problems) SDQ scores, and on teacher reported reactive aggression. Meta-analytic reviews of psycho-social interventions [70] and parent-training specifically [73] have shown that treatment effect sizes on teacher-reported outcomes are small (.13 and .15 respectively). It is not possible to infer from our findings with this modest sample size that RICAP did <u>not</u> lead to comparable effects on teacher rated outcomes, however it is possible that RICAP has an effect which is confined to behaviours shown within the family. In particular, RICAP is an intervention focussed on improving the parent child relationship, addressing attachment related concerns and enhancing reflection on the meaning of behaviours whether these might reflect cognitive limitations or appraisals, emotional distress or limitations in emotional expression, social or motivational needs, defensive processes or threat related concerns. As the focus is on creating change in the home setting this may not generalise to the school setting. In future, RICAP could be expanded to include a ‘relationships in school’ element aiming to increase reflective functioning in that setting ideally including sessions with teacher and child. A school-based version of the Incredible Years programme has been developed and shown to be associated with a small effect on teacher-reported outcomes [74]). In contrast to the RICAP arm, the CTAU in this study did include some school focused content for 42% of children which may explain the significant treatment effect on SDQ total problems in the CTAU arm.

The lack of group differences on parent and teacher SDQ conduct problems scores may reflect that it was designed as an epidemiological screening tool, and its small number of items may make it less sensitive to change than the CBCL. In addition, two out of five conduct problem items (lying and stealing) are less relevant for younger children. Smaller effect sizes for the SDQ compared to the CBCL or the Eyberg Child Behaviour Inventory, another lengthier measure, have been found in previous RCTs [3,75].

Strengths of the study include that it was a pragmatic trial [76], run within community settings, employing regular clinical staff, and it did not exclude cases on the basis of comorbidity. These design elements enhance the generalisability of the findings from the study. The extent to which trial outcomes reported are ecologically valid is important [77] and RCTs are hard to complete in real-life settings. One possible source of bias in pragmatic trials can be clinician de-selection of vulnerable children. To address this the study used consecutive referrals to parenting groups which meant that this was very unlikely. The initial rate of consent to take part in the study was high and very few eligible families refused randomisation; one declined in order to choose child psychotherapy and 3 families did not want further intervention after PMT. One child was admitted to an inpatient unit by his clinician.

A major limitation of the study arose from the finding of superiority for parent-reported but not teacher-reported outcomes. Parents were not blinded to treatment, and parent report may have simply reflected greater satisfaction among parents for RICAP than for CTAU. Teachers by contrast had not received either intervention and were blind to the intervention status of the child. A further limitation is that both CTAU and RICAP followed PMT and it is therefore not clear whether their effectiveness was potentiated by this first intervention. Interpretation of the PDR findings is limited by smaller numbers of parents completing this measure than the others, 20/28 in the RICAP group and 25/30 in CTAU which could have introduced unknown biases in the subsample providing PDR data. The smaller number of parents reporting suggests that this measure is less feasible than others for use in trials. We have been unable to locate any RCTs which used both the CBCL and the PDR, so it remains unclear whether it is a valid measure of treatment effect. The sample was overwhelmingly white British so the findings may not generalise to other ethnic groups.

Although the first phase of the study was uncontrolled, so direct comparison with published trials cannot be made, there were indications that numbers still in the clinical range following IY were higher than in other studies [3]. We suggest three possible explanations. First our children were considered to be in the clinical range following IY if they were above threshold either on the CBCL or SDQ, either by parent or teacher report. In view of the importance of conduct problems outside of the family to peer and educational development we regard this as a strength, which also likely lowered the threshold for selection into the trial. Second, again in contrast to Scott [3], adherence to IY was not monitored which may have affected the effectiveness of the intervention, and third there was a higher drop-out from IY, of 40% in this study compared to 19% in the study of Scott [3].

In conclusion, this study demonstrated the feasibility of a design to evaluate a new intervention for the clearest target that we know of for early intervention to improve the mental health of children, adolescents and adults. It introduces a key step towards personalisation based on lack of responsiveness to a first line treatment for the disorder [78]. The new RICAP intervention was highly acceptable to parents and children, and addresses a plea made by the parents of children who are offered parent training, that assessments and treatments should focus as much on the child as on the parents [33].

Based on the findings there is a need for a full-scale RCT of RICAP, with sufficient statistical power to examine both parent and teacher reported outcomes. Many of the ideas and techniques in RICAP, which focuses on the child and the family, are well suited to adaptations to a focus on the child and their peers, and the child in the classroom, and hence potentially to an approach tailored to the particular social contexts of the child’s behaviours.

## Data Availability

The participants of this study did not give written consent for their data to be shared publicly, so due to the sensitive nature of the research supporting data is not available.

## Acknowledgements

We thank all the children and families and teachers for their participation; the service managers and multi-professional therapists from within CAMHS services on Wirral (Child and Family Service, Cheshire and Wirral Partnership NHS Foundation Trust) and in Liverpool (Child and Family, Royal Liverpool Children’s NHS Trust) including multiagency BEST therapists for supporting the study throughout; Bron Milligan, Dr Mary Naughton, Elspeth Bromiley and Jo Pearce as lead clinicians for parenting groups; Dr Gillian Lancaster for ensuring an independent randomisation procedure; Prof Richard Harrington for advice on study design and evaluation in the early planning phases.

