## Supplementary material for "Toward improving outcomes from early intervention for vulnerable children: pilot RCT of a new treatment for non-responders to parent training for conduct problems"

**Supplementary Materials 1:** Calculation of sample size

The study design anticipated that 160 families would be needed in phase 1 in order to generate a total of 60 for randomization at time 2. This was based on an attrition rate of 25% and a prediction that around 50% of the children would remain in the clinical range following parent training. The sample size for randomisation was based on Browne’s (1995) recommendation that in a pilot study 30 participants are needed in each group to provide reasonably narrow confidence intervals for the standard deviations required for a power calculation.

**Supplementary Materials 2:** Information given to participants

In order to minimise expectancy effects parents were told at the point of consent before phase 1 intervention that the study was designed to compare two different ways of working with children and parents after parent training. The information sheets were worded carefully. The study was entitled ‘Matching therapy to children’s needs: a comparison of different ways of helping’.

Parents were told:

***What is the study about?***

*Research shows that most children are helped when their parents participate in parenting groups like the one you have been invited to join. Some children also seem to benefit from having their own additional time with a therapist. However, research does not yet tell us which is the best way to give this additional help. That is what we want to find out. The parenting group you have been invited to join lasts for between 12 and 14 weeks. If after you have attended the parenting group, you, or your child’s teacher, tell us in a questionnaire that your child is showing behaviour problems above a certain level, you will then be offered one of two possible types of further help.*

***Why do we need to compare two types of help?***

*We want and need to know which is the better way of working with children. In one method, therapists have a set step-by-step plan and work through this with you and your child. In the other, the therapist works through the difficulties you and your child want to talk about each time you meet. Both of the methods have been used before and are still being used today to help children. But we still do not know which is the better type of help. The results of this research study should hopefully give us an answer.*

**Table S1** (a) Baseline (time 1) and time 2 informant report of child symptoms pre and post phase 1 intervention for whole sample and (b) time 1 and time 2 informant report of child symptoms for subgroups who were randomly allocated to each phase 2 treatment, those who were not eligible for phase 2 treatment and those eligible who were not randomised.

|  | (a) Phase 1 intervention | | (b) Subgroups defined by eligibility for random allocation to phase 2 intervention;  mean (SD) pre and post phase 1 intervention | | | | | | | |
| --- | --- | --- | --- | --- | --- | --- | --- | --- | --- | --- |
|  | Whole sample; mean (SD)  (N=102) | | Not eligible after phase 1 treatment (n=28) | | Eligible but not randomised (n=12) | | RICAP  (n=31) | | TAU  (n=31) | |
|  | Time 1 | Time 2 | Time 1 | Time 2 | Time 1 | Time 2 | Time 1 | Time 2 | Time 1 | Time 2 |
| % PMT sessions  attended | 55.6  (32.1) | _ | 61.0  (29.8) | _ | 48.4  (31.1) | _ | 50.9  (32.5) | _ | 58.2  (34.4) | _ |
| Parent  CBCL externalising | 28.7  (12.2) | 23.5 **  (13.1) | 17.3  (8.4) | 10.4  (5.1) | 33.3  (10.6) | 26.7  (11.9) | 33.9  (11.3) | 30.1  (11.0) | 32.74  (8.8) | 28.80  (11.8) |
| SDQ conduct | 5.7  (2.4) | 4.4 **  (2.4) | 4.0  (1.8) | 2.1  (1.0) | 6.17  (2.7) | 4.73  (2.0) | 6.71  (2.1) | 5.39  (2.0) | 6.19  (2.2) | 5.63  (2.4) |
| Teacher  CBCL externalising | 17.3  (15.7) |  | 8.2  (8.7) | 6.0  (6.0) | 23.8  (17.9) | 18.6  (17.0) | 21.6  (18.8) | 18.2  (16.7) | 19.31  (13.4) | 19.93  (14.3) |

**Table S1** (continued).

|  | (a) Phase 1 intervention | | (b) Subgroups defined by eligibility for random allocation to phase 2 intervention;  mean (SD) pre and post phase 1 intervention | | | | | | | |
| --- | --- | --- | --- | --- | --- | --- | --- | --- | --- | --- |
|  | Whole sample; mean (SD)  (N=102) | | Not eligible after phase 1 treatment (n=28) | | Eligible but not randomised (n=12) | | RICAP  (n=31) | | TAU  (n=31) | |
|  | Time 1 | Time 2 | Time 1 | Time 2 | Time 1 | Time 2 | Time 1 | Time 2 | Time 1 | Time 2 |
| SDQ conduct | 2.6  (2.5) | 2.3 *  (2.3) | 1.3  (1.7) | 1.0  (1.2) | 3.0  (2.3) | 2.1  (1.8) | 3.3  (3.1) | 2.7  (2.8) | 3.03  (2.2) | 3.14  (2.6) |

*, ** Paired t-test assessing whole group change pre (time 1) to post phase 1 (time 2) parent management training intervention indicated significant reduction in symptom reports at P < 0.05; P < 0.001 level respectively.

**Table S2** Clinically reliable change following phase 2 intervention: Proportion of cases within clinical, borderline and normal ranges on parent report measures at each time point for children in the RICAP and TAU treatment arms.

| Measure | Intervention | Assessment Time | Proportion (%) scoring in each score range on scale | | | | | | |
| --- | --- | --- | --- | --- | --- | --- | --- | --- | --- |
|  |  |  | Parent report | | | | Teacher report | | |
|  |  |  | Normal | Borderline | Clinical |  | Normal | Borderline | Clinical |
| SDQ  Conduct | RICAP | Before  After | 3.2  3.6 | 9.7  21.4 | 87.1  75.0 |  | 60.0  55.6 | 10.0  7.4 | 30.0  37.0 |
|  | CTAU | Before  After | 10.0  13.7 | 6.7  13.8 | 83.3  72.5 |  | 37.9  57.1 | 17.2  14.3 | 44.8  28.6 |
| SDQ total problems | RICAP | Before  After | 6.5  17.9 | 25.8  21.4 | 67.7  60.7 |  | 50.0  33.3 | 16.7  25.9 | 33.3  40.7 |
|  | CTAU | Before  After | 13.3  31.0 | 13.3  10.3 | 73.3  58.6 |  | 41.4  64.3 | 10.3  10.7 | 48.3  25.0 |
| CBCL externalising | RICAP | Before  After | 6.5  32.1 | 6.5  10.7 | 87.1  57.1 |  | 50.0  40.7 | 6.7  22.2 | 43.3  37.0 |

**Table S2** (continued).

| Measure | Intervention | | Assessment Time | | Proportion (%) scoring in each score range on scale | | | | | | | | | | | | | |
| --- | --- | --- | --- | --- | --- | --- | --- | --- | --- | --- | --- | --- | --- | --- | --- | --- | --- | --- |
|  |  |  |  |  | Parent report | | | | | | | | Teacher report | | | | | |
|  |  |  |  |  | Normal | | Borderline | | Clinical | |  | | Normal | | Borderline | | Clinical | |
| CBCL externalising | RICAP | Before  After | | 6.5  32.1 | | 6.5  10.7 | | 87.1  57.1 | |  | | 50.0  40.7 | |  |  |  |  |  |
| CBCL externalising | | CTAU | | Before  After | | 13.3  17.2 | | 6.7  6.9 | | 80.0  75.9 | |  | | 37.9  46.4 | | 13.8  14.3 | | 48.3  39.3 |
| CBCL internalising | | RICAP | | Before  After | | 35.5  60.7 | | 16.1  17.9 | | 48.4  21.4 | |  | | 73.3  55.6 | | 10.0  11.1 | | 16.7  33.3 |
|  | | CTAU | | Before  After | | 36.7  34.5 | | 26.7  13.8 | | 36.7  51.7 | |  | | 69.0  78.6 | | 24.1  3.6 | | 6.9  17.9 |
